# Echocardiographic characterization and markers of cardiovascular risk in adults with sickle cell disease in a Colombian tertiary referral centre: a cross-sectional study

**DOI:** 10.64898/2026.04.16.26351071

**Authors:** Martín E. Arrieta-Mendoza, Juan R. Betancourt, Sebastián Ayala-Zapata, Stephany Barbosa-Balaguera, Christian D. Messu-Llanos, Juan P. Rosales-Melo, Darío F. Andrade-Hoyos, Álvaro Herrera-Escandón, Oswaldo E. Aguilar-Molina

## Abstract

Sickle cell disease (SCD) is associated with substantial cardiovascular morbidity, but echocardiographic data from Latin American populations remain scarce. We aimed to characterise the structural, functional, and haemodynamic echocardiographic profile of adults with SCD attending a tertiary referral centre in Cali, Colombia. We conducted an observational, cross-sectional study based on systematic review of medical records and transthoracic echocardiography reports of consecutive adult patients (≥18 years) with confirmed SCD evaluated between January 2022 and December 2024. Patients with complex congenital heart disease, severe valvular disease of unrelated aetiology, pregnancy, or echocardiograms of insufficient quality were excluded. Of 669 patients screened, 57 met inclusion criteria. Reporting followed STROBE recommendations. The median age was 24 years (interquartile range [IQR] 21–32) and 59.6% were female; the SS genotype was the most frequent (76.4%) and 71.4% were on hydroxyurea. Median haemoglobin was 10.2 g/dL (IQR 9.3–11.4) and median NT-proBNP 491 pg/mL (IQR 98–1290). Most patients had preserved left ventricular dimensions and systolic function (median ejection fraction 63%, IQR 57–66.5; mean global longitudinal strain −18.9% ± 2.9). Right ventricular function was preserved (mean tricuspid annular plane systolic excursion 25.4 ± 4.6 mm). Left ventricular geometry was normal in 42.1%, with concentric remodelling in 24.6%, concentric hypertrophy in 21.1%, and eccentric hypertrophy in 12.3%. Diastolic function was normal in 71.4%. Valvular disease, when present, was predominantly mild. Tricuspid regurgitation velocity exceeded 2.5 m/s in 29.8% of patients and exceeded 3.0 m/s in 10.5%, identifying a substantial subgroup at intermediate-to-high probability of pulmonary hypertension. In this Colombian cohort of relatively young adults with SCD, cardiac structure and biventricular function were largely preserved, but nearly one-third of patients had echocardiographic findings suggestive of pulmonary hypertension. These findings support the routine use of transthoracic echocardiography as an accessible tool for early cardiovascular risk stratification in adults with SCD in low- and middle-income settings.

## Introduction

Sickle cell disease (SCD) is an inherited haemoglobinopathy caused by a single point mutation in the *HBB* gene that produces an abnormal β-globin chain. The resulting haemoglobin S polymerises under hypoxic conditions, deforming erythrocytes and triggering chronic haemolysis, recurrent vaso-occlusion, and progressive multi-organ damage [1,2]. Globally, more than 7.7 million people are estimated to live with SCD, with the highest burden concentrated in sub-Saharan Africa and Latin America [3]. In Colombia, the national prevalence is approximately 0.33 cases per 100,000 inhabitants, but rises to 0.66 per 100,000 in Valle del Cauca, where Afro-descendant ancestry is highly represented [4].

Improvements in early diagnosis, infection prophylaxis, and disease-modifying therapies—particularly hydroxyurea—have substantially extended survival in SCD over the last three decades. As a consequence, cardiopulmonary complications have emerged as the leading cause of premature death in adults with SCD, accounting for 26–43% of mortality in published cohorts [5,6]. Reported manifestations include left ventricular hypertrophy and chamber dilatation, diastolic dysfunction, pulmonary hypertension, valvular regurgitation, atrial arrhythmias, and—less commonly—overt systolic dysfunction [7–9]. Tricuspid regurgitation velocity (TRV) >2.5 m/s, an E/e′ ratio >10, and impaired global longitudinal strain (GLS) have all been associated with increased mortality in this population [8,10]. Transthoracic echocardiography (TTE) remains the cornerstone non-invasive imaging modality for cardiovascular assessment in SCD: it is widely available, inexpensive, free of ionising radiation, and capable of detecting subclinical structural and functional alterations long before symptoms develop [9,11].

Despite the relevance of cardiovascular involvement in SCD, echocardiographic data from Latin American populations are very limited. Most published cohorts originate from North America, Europe, the Middle East, or sub-Saharan Africa [8,11–14], and direct extrapolation to populations with different genetic background, age structure, comorbidity profile, and access to disease-modifying therapy is questionable. To our knowledge, no contemporary cohort has systematically described the echocardiographic profile of adults with SCD in the Colombian Pacific—a region with one of the highest concentrations of Afro-descendant population in the country.

The aim of this study was to characterise the clinical and echocardiographic features of adults with SCD attending a tertiary referral centre in Cali, Colombia, with particular focus on structural, systolic, diastolic, valvular, and haemodynamic parameters relevant to cardiovascular risk stratification.

## Methods

### Study design and reporting

We conducted an observational, descriptive, cross-sectional study based on systematic review of electronic medical records and TTE reports of consecutive adult patients with a confirmed diagnosis of SCD evaluated at the Hospital Universitario del Valle “Evaristo García” E.S.E. (HUV), a tertiary referral centre in Cali, Colombia, between January 2022 and December 2024. Reporting follows the Strengthening the Reporting of Observational Studies in Epidemiology (STROBE) recommendations [15]; a completed STROBE checklist is provided as Supporting Information (S1 Checklist).

### Setting and participants

The HUV is the largest public tertiary hospital in southwestern Colombia and serves as the regional referral centre for haemoglobinopathies in Valle del Cauca and adjacent departments of the Pacific region.

#### Inclusion criteria

(i) age ≥18 years; (ii) confirmed diagnosis of SCD established by haemoglobin electrophoresis or DNA-based methods; and (iii) at least one complete TTE performed and reported during the study period.

#### Exclusion criteria

(i) complex structural congenital heart disease unrelated to SCD; (ii) severe valvular heart disease of aetiology unrelated to SCD; (iii) incomplete TTE studies or studies of insufficient image quality for analysis; (iv) inability to verify clinical or genetic confirmation of SCD; and (v) pregnancy at the time of TTE.

### Data collection and definitions

Demographic, clinical, laboratory, and echocardiographic variables were extracted using a standardised electronic data collection form with double data entry to minimise transcription errors. Echocardiographic measurements were obtained from validated reports issued by board-certified cardiologists with level III training in echocardiography, performed on commercially available ultrasound systems and reported in accordance with the recommendations of the American Society of Echocardiography (ASE) and the European Association of Cardiovascular Imaging (EACVI) [16].

Left ventricular geometry was classified using relative wall thickness and indexed left ventricular mass per ASE/EACVI criteria [16]. Diastolic function was graded using the 2016 ASE/EACVI algorithm [17]. The probability of pulmonary hypertension was assessed using the European Society of Cardiology/European Respiratory Society echocardiographic criteria, with TRV >2.5 m/s and >2.8 m/s used as thresholds for intermediate and high probability, respectively [18]. Global longitudinal strain (GLS) was measured by two-dimensional speckle-tracking echocardiography in vendor-specific software and is reported as the absolute mean value with standard deviation (SD).

### Statistical analysis

Continuous variables were tested for normality using the Shapiro–Wilk test and visual inspection of histograms; they are reported as mean ± SD when normally distributed and as median with interquartile range (IQR) otherwise. Categorical variables are reported as absolute frequencies and percentages. The analysis was descriptive, in line with the cross-sectional design and pre-defined objectives. Missing data were not imputed and the denominator is reported for each variable. Statistical analyses were performed using Stata version 18 (StataCorp LLC, College Station, TX, USA).

## Ethics

The study was approved by the Research Ethics Committee of the Universidad del Valle (approval reference available on request). The requirement for individual informed consent was waived in accordance with national regulations for retrospective research with anonymised data (Resolution 8430 of 1993, Ministry of Health of Colombia), which classifies this study as risk-free research. All procedures were conducted in accordance with the Declaration of Helsinki. Patient data were anonymised before extraction and analysis.

## Results

### Study population

Of 669 adult records initially screened from the hospital information systems, 612 were excluded according to the criteria detailed in **Fig 1**, leaving 57 patients in the final analysis.

**Fig 1.**
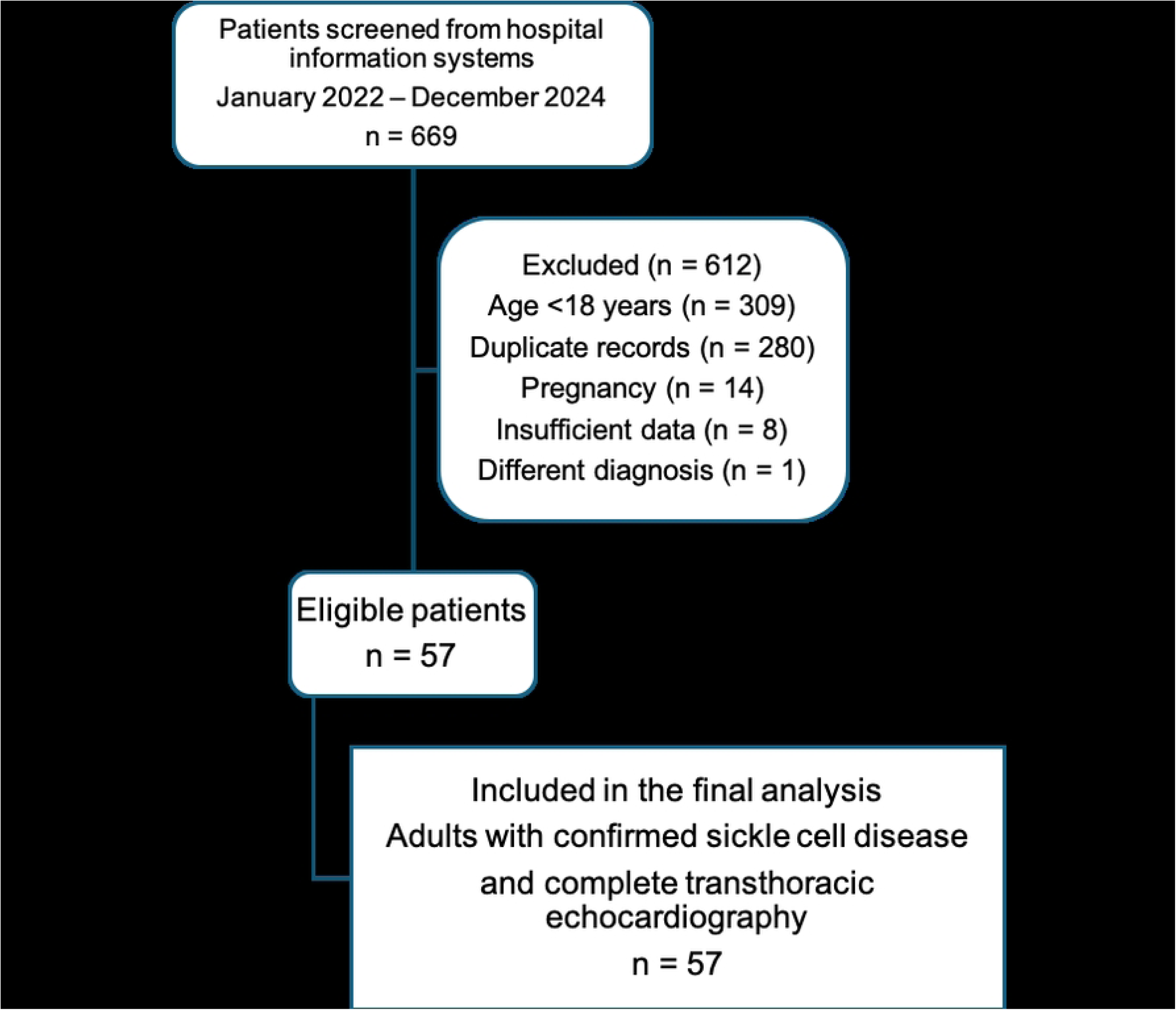
Study flow diagram. Of 669 patients screened from hospital information systems between January 2022 and December 2024, 612 were excluded (309 aged <18 years, 280 duplicate records, 14 pregnant, 8 with insufficient data, 1 with a different diagnosis), leaving 57 adults with confirmed sickle cell disease and complete transthoracic echocardiography for the final analysis.

Baseline clinical and laboratory characteristics are summarised in **Table 1**. The median age was 24 years (IQR 21–32), and 34 patients (59.6%) were female. The SS genotype was the most frequent, present in 42 patients (76.4%); SC, Sβ^+^, Sβ^0^, and other genotypes accounted for the remainder. A vaso-occlusive crisis within the previous 30 days was reported in 37 patients (66.1%), and 40 (71.4%) were receiving hydroxyurea at the time of evaluation. Median haemoglobin was 10.2 g/dL (IQR 9.3–11.4); median creatinine 0.5 mg/dL (IQR 0.4–0.6); median NT-proBNP 491 pg/mL (IQR 98–1290). New York Heart Association (NYHA) functional class distribution is shown in **Fig 2**: 84% of patients were in class I, 10% in class II, and 6% in class IV.

**Fig 2.**
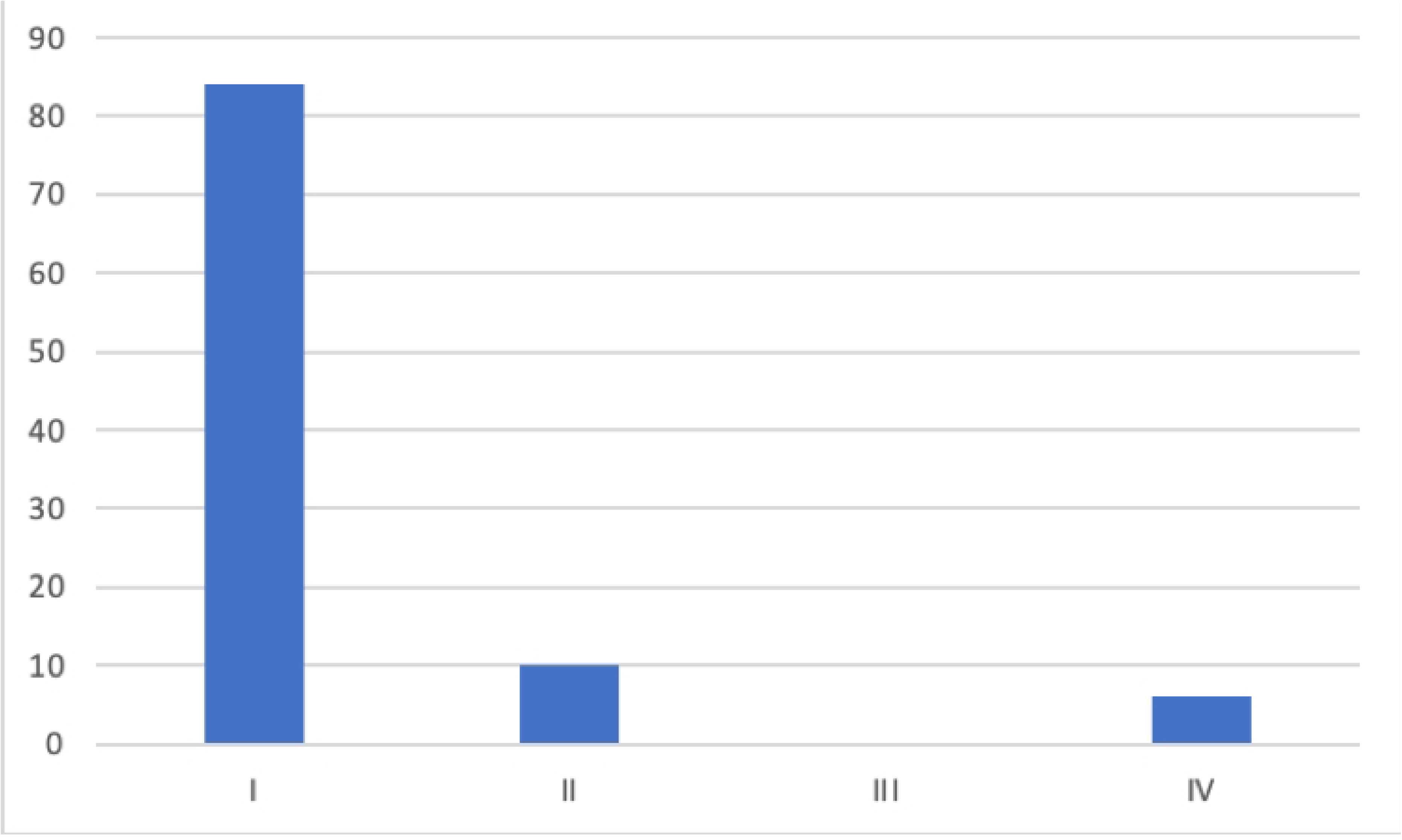
Distribution of New York Heart Association (NYHA) functional class. Bar chart showing the proportion of patients in each NYHA class. NYHA: I, no limitation of physical activity; II, slight limitation; III, marked limitation; IV, symptoms at rest.

**Table 1.**
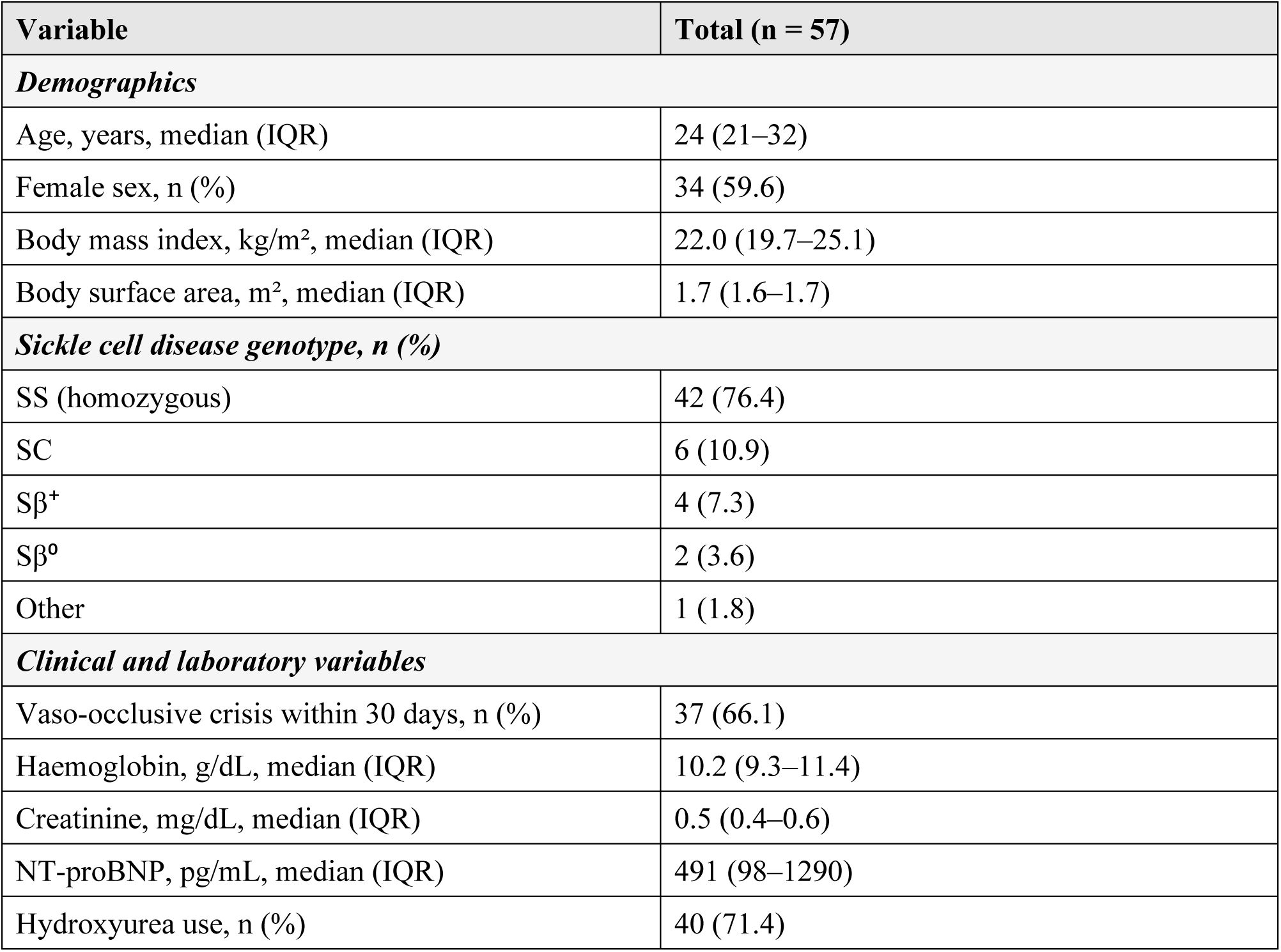
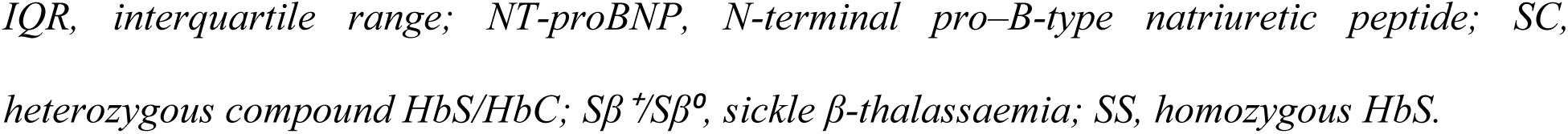
Baseline clinical and laboratory characteristics of the study population.

| Variable | Total (n = 57) |
| --- | --- |
| ***Demographics*** |  |
| Age, years, median (IQR) | 24 (21–32) |
| Female sex, n (%) | 34 (59.6) |
| Body mass index, kg/m<sup>2</sup>, median (IQR) | 22.0 (19.7–25.1) |
| Body surface area, m<sup>2</sup>, median (IQR) | 1.7 (1.6–1.7) |
| ***Sickle cell disease genotype, n (%)*** |  |
| SS (homozygous) | 42 (76.4) |
| SC | 6 (10.9) |
| Sβ<sup>+</sup> | 4 (7.3) |
| Sβ<sup>o</sup> | 2 (3.6) |
| Other | 1 (1.8) |
| ***Clinical and laboratory variables*** |  |
| Vaso-occlusive crisis within 30 days, n (%) | 37 (66.1) |
| Haemoglobin, g/dL, median (IQR) | 10.2 (9.3–11.4) |
| Creatinine, mg/dL, median (IQR) | 0.5 (0.4–0.6) |
| NT-proBNP, pg/mL, median (IQR) | 491 (98–1290) |
| Hydroxyurea use, n (%) | 40 (71.4) |

*IQR, interquartile range; NT-proBNP, N-terminal pro-B-type natriuretic peptide; SC, heterozygous compound HbS/HbC; Sβ <sup>+</sup>/Sβ <sup>0</sup>, sickle $\beta$ -thalassaemia; SS, homozygous HbS.*

### Structural and systolic function

Echocardiographic measurements are summarised in **Table 2**. Left ventricular dimensions, wall thickness, and indexed mass were within normal reference ranges in most patients. Left ventricular geometry was normal in 24 patients (42.1%); concentric remodelling was observed in 14 (24.6%), concentric hypertrophy in 12 (21.1%), and eccentric hypertrophy in 7 (12.3%). Median left ventricular ejection fraction was 63% (IQR 57.0–66.5), and mean GLS was −18.9% ± 2.9, both within reference ranges. Right ventricular systolic function was preserved (mean tricuspid annular plane systolic excursion 25.4 mm ± 4.6; median tricuspid annulus systolic velocity [S′] 15 cm/s, IQR 13–16).

**Table 2.**
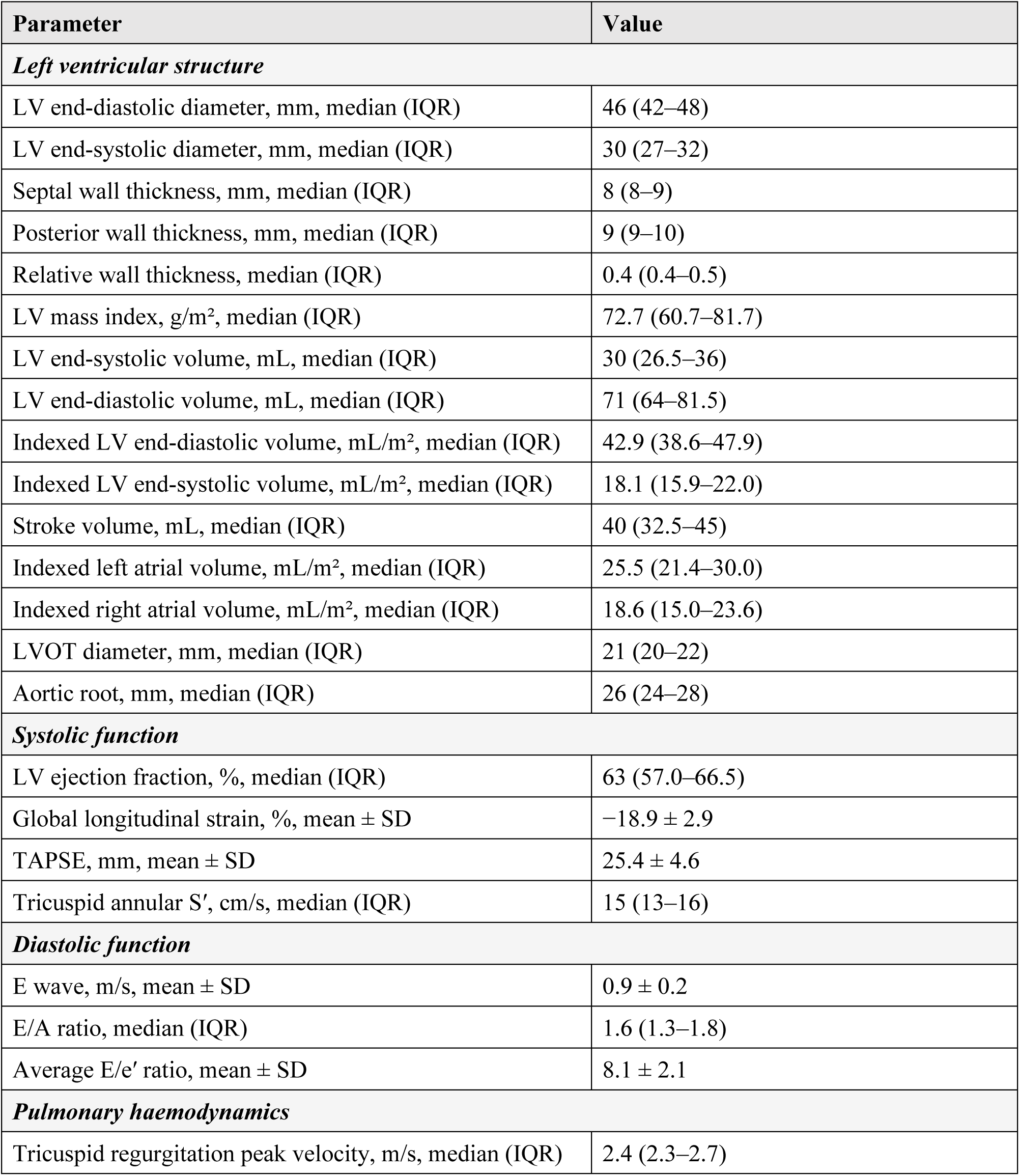
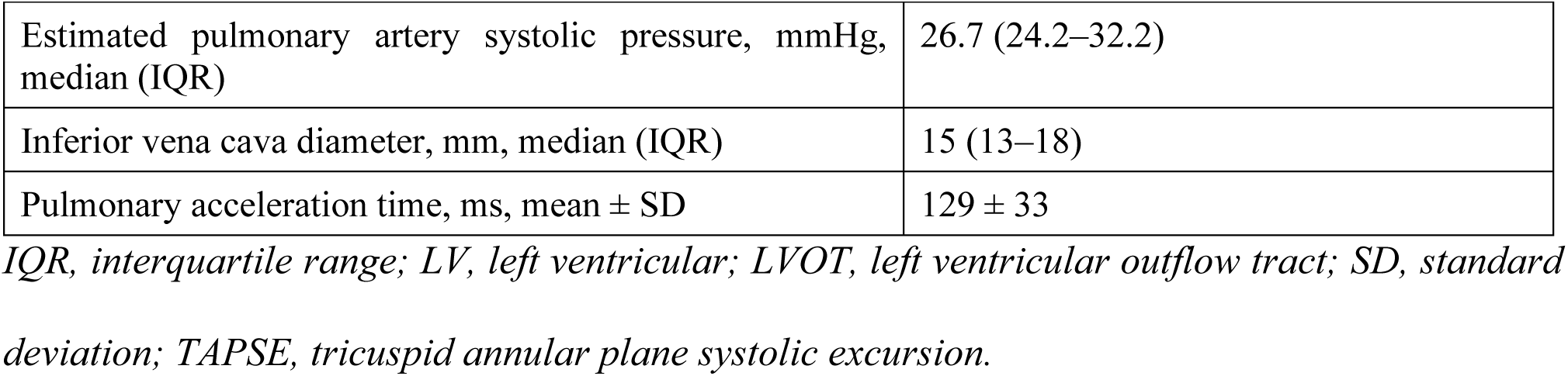
Echocardiographic measurements of the study population (n = 57).

### Valvular and diastolic function

Valvular abnormalities, when present, were predominantly mild (**Fig 3**). Mild tricuspid regurgitation was the most frequent finding (≈86%), followed by mild mitral regurgitation (≈72%) and mild aortic regurgitation (≈18%). Moderate tricuspid regurgitation was observed in approximately 7% of patients and severe tricuspid regurgitation in approximately 2%.

**Fig 3.**
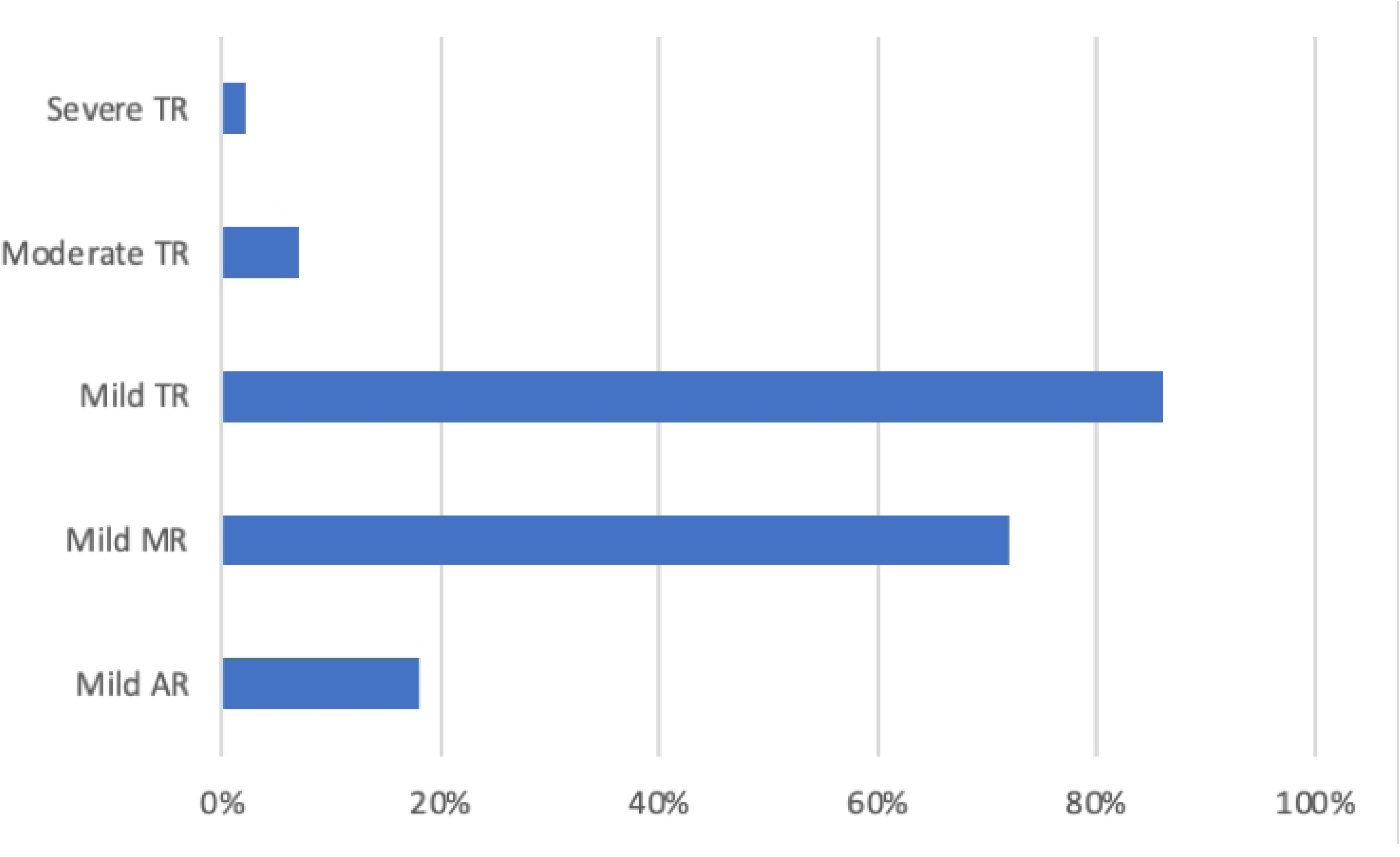
Prevalence of valvular regurgitation. Horizontal bar chart showing the proportion of patients with mild, moderate, or severe regurgitation of the aortic, mitral, and tricuspid valves. AR, aortic regurgitation; MR, mitral regurgitation; TR, tricuspid regurgitation.

Diastolic function was normal in 41 patients (71.4%); grade I (impaired relaxation) was present in 12 (21.4%) and grade II in 4 (7.1%); no patient had grade III diastolic dysfunction (**Fig 4**). The median E/A ratio was 1.6 (IQR 1.3–1.8), and the mean E/e′ ratio was 8.1 ± 2.1. The median indexed left atrial volume was 25.5 mL/m² (IQR 21.4–30.0), within normal limits.

**Fig 4.**
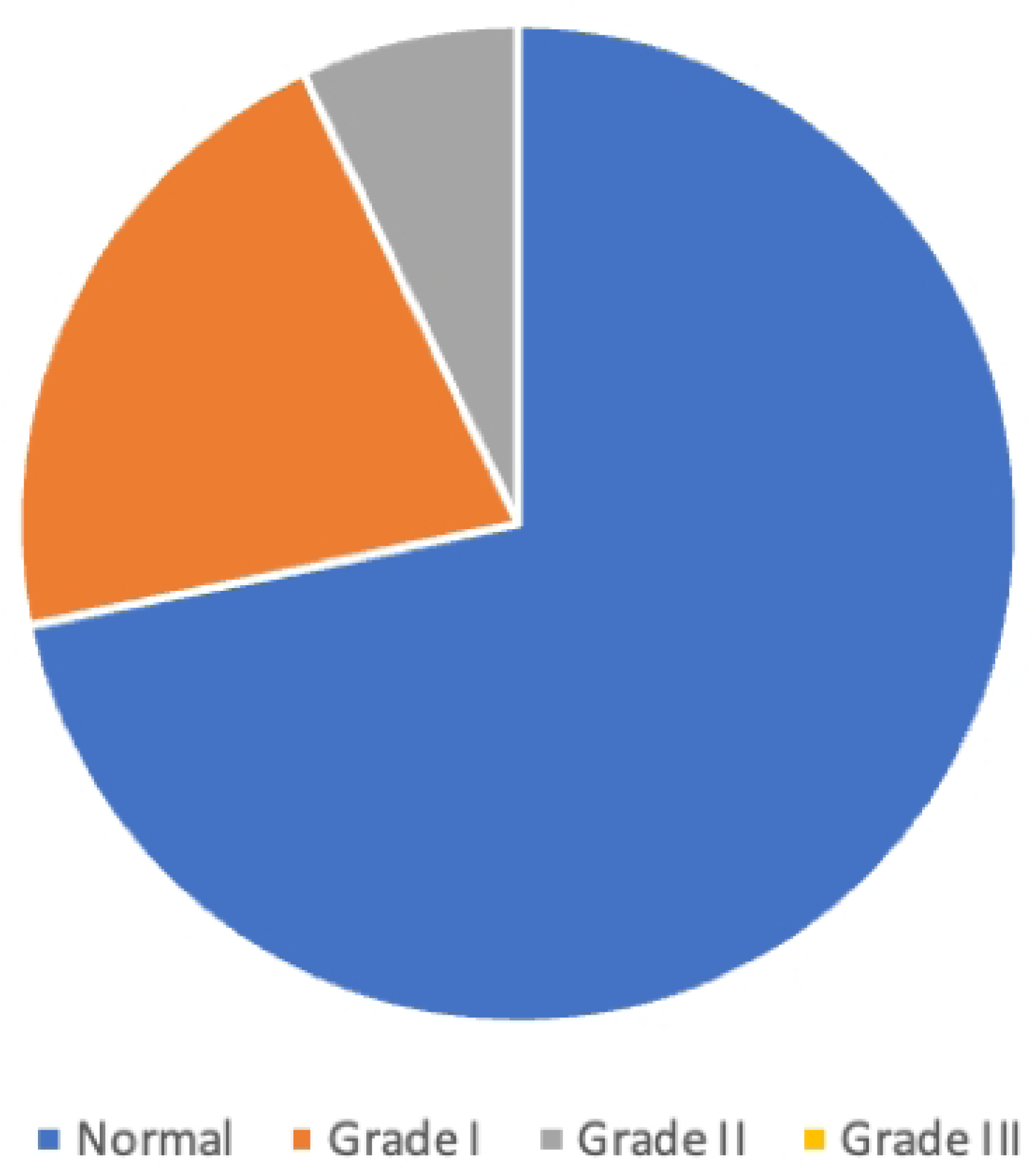
Distribution of diastolic function. Pie chart showing the proportion of patients with normal diastolic function and with grades I, II, and III diastolic dysfunction.

### Pulmonary haemodynamics

Median TRV was 2.4 m/s (IQR 2.3–2.7). Seventeen patients (29.8%) had a TRV >2.5 m/s, and 6 (10.5%) had a TRV >3.0 m/s. The median estimated pulmonary artery systolic pressure was 26.7 mmHg (IQR 24.2–32.2), the median inferior vena cava diameter was 15 mm (IQR 13–18), and the mean pulmonary acceleration time was 129 ± 33 ms.

## Discussion

In this contemporary cross-sectional study of 57 adults with SCD attending a tertiary referral centre in Cali, Colombia, we found that cardiac structure and biventricular systolic function were largely preserved at the population level, while approximately one-third of patients had echocardiographic findings consistent with an intermediate-to-high probability of pulmonary hypertension. To our knowledge, this is the first systematic echocardiographic characterisation of adults with SCD in the Colombian Pacific, a region with one of the highest concentrations of Afro-descendant population in Latin America.

Our cohort was younger (median age 24 years) than most published series, in which median ages typically range between 32 and 37 years [8,11,19]. Two factors may explain this difference. First, our centre acts as a regional referral hub for both paediatric and adult haemoglobinopathy care, with continuous transition of younger patients into adult clinics. Second, premature mortality in SCD remains high in low- and middle-income settings, which truncates the age distribution of prevalent adult cases [3,5]. This younger age structure has direct implications for the interpretation of our findings, as both chamber remodelling and pulmonary hypertension increase with cumulative disease exposure [8,11].

The female predominance (59.6%) in our sample is comparable to that reported by Parent et al. [8] and Naoman et al. [13]. Because SCD is an autosomal recessive disorder, equal sex distribution is biologically expected; the observed predominance is most likely explained by greater contact of women with health services and referral patterns rather than a true epidemiological difference.

Ventricular and atrial volumes—both absolute and indexed to body surface area—were within normal reference limits in the majority of patients, in contrast with the cohort reported by Abdul-Mohsen [11], in which up to one-quarter of patients had chamber dilatation, and more consistent with younger Western series [12,19]. Indexed left ventricular mass was likewise preserved. Two factors plausibly account for these results: the relatively young age of our cohort (with a shorter cumulative duration of chronic anaemia and volume overload) and the high proportion of patients on hydroxyurea (71.4%), which has been shown to reduce haemolysis and may attenuate adverse cardiac remodelling [20].

Left ventricular systolic function was preserved at the population level (median ejection fraction 63%; mean GLS −18.9%), in agreement with previous series [11,12,16,19]. The mean GLS value falls within the normal range reported for healthy adults of similar age and is comparable to data from European cohorts of patients with SCD [12]. Right ventricular longitudinal function was also preserved, in contrast with the report by Chiadika et al. [14] in which approximately one-third of patients had abnormal tricuspid annular plane systolic excursion. Differences in age structure and disease burden are the most likely explanation.

Diastolic function was normal in most patients (71.4%), and no patient had grade III dysfunction. This contrasts with cohorts in which the majority of patients showed some degree of diastolic dysfunction [17] but is consistent with the Saudi cohort reported by Abdul-Mohsen [11], in which dysfunction—when present—was predominantly mild. Diastolic dysfunction has been independently associated with mortality in SCD [19]; the relatively low prevalence in our cohort may reflect both younger age and high hydroxyurea use, but mortality outcomes could not be evaluated in this cross-sectional design.

The most clinically relevant finding of our study is that 29.8% of patients had a TRV >2.5 m/s, and 10.5% had a TRV >3.0 m/s, identifying a substantial subgroup at intermediate-to-high echocardiographic probability of pulmonary hypertension [18]. These proportions are remarkably consistent with the landmark French cohort reported by Parent et al. [8] (TRV >2.5 m/s in approximately 27%) and with North American series [21,22], suggesting that the burden of pulmonary hypertension risk in our Colombian cohort is comparable to that reported elsewhere despite the younger median age. TRV >2.5 m/s has been shown to confer a more than ten-fold increase in mortality risk in SCD when combined with other markers such as elevated E/e′ ratio [10]; identifying these patients echocardiographically may therefore guide earlier referral for confirmatory right heart catheterisation and intensification of disease-modifying therapy [9].

From a public-health perspective, our data support the routine use of TTE as an accessible, non-invasive tool for cardiovascular risk stratification in adults with SCD in low- and middle-income settings, where access to advanced cardiovascular imaging modalities is often limited. Identification of an intermediate-to-high probability of pulmonary hypertension on TTE should prompt further evaluation and consideration of early intensification of disease-modifying therapy, rather than waiting for symptomatic deterioration.

### Strengths and limitations

#### Strengths

This is, to our knowledge, the first systematic echocardiographic characterisation of adults with SCD in the Colombian Pacific. All echocardiograms were performed and reported by board-certified cardiologists with level III training using a standardised institutional protocol, and we report a comprehensive set of structural, systolic, diastolic, valvular, and haemodynamic parameters, including GLS by speckle-tracking echocardiography.

#### Limitations

Our study has several limitations that must be acknowledged. First, the retrospective single-centre design limits external generalisability and precludes causal inference. Second, the absence of a healthy local control group prevents direct comparison of echocardiographic parameters with reference values derived from the same population. Third, the modest sample size (n = 57) and the descriptive nature of the analysis precluded multivariable modelling of predictors of pulmonary hypertension or diastolic dysfunction. Fourth, mortality and longitudinal cardiovascular outcomes were not evaluated. Fifth, right heart catheterisation was not systematically performed; pulmonary hypertension probability is therefore based on echocardiographic criteria alone. Finally, the relatively young age of the cohort may have underestimated the burden of cardiovascular involvement that typically accumulates beyond the fifth decade of life. These limitations should be addressed in future prospective multicentre studies including longitudinal follow-up and right heart catheterisation in patients with elevated TRV.

## Conclusion

In this Colombian cohort of relatively young adults with SCD, cardiac structure and biventricular systolic and diastolic function were largely preserved. Nonetheless, nearly one-third of patients had echocardiographic findings consistent with an intermediate-to-high probability of pulmonary hypertension, identifying a clinically relevant subgroup at increased cardiovascular risk. Our findings support the use of transthoracic echocardiography as a routine, accessible tool for early cardiovascular risk stratification in adults with SCD in Latin American and other low- and middle-income settings, and provide a contemporary local reference against which future prospective and interventional studies can be designed.

## Data Availability

All relevant data are within the manuscript, its tables, and its Supporting Information files. The de-identified individual-level dataset used for the analyses is provided as Supporting Information (S2 Dataset).

## Acknowledgments

The authors thank the staff of the Department of Cardiology and the Echocardiography Laboratory of the Hospital Universitario del Valle “Evaristo García” E.S.E. for their support during data retrieval.

## Author contributions

**Conceptualization:** MEAM, JRB, AHE.

**Data curation:** MEAM, JRB, CDML, JPRM, DFAH.

**Formal analysis:** SAZ, SBB, MEAM, OEAM.

**Investigation:** MEAM, JRB, CDML, JPRM, DFAH, SAZ.

**Methodology:** MEAM, AHE, SAZ, SBB.

**Supervision:** AHE, OEAM.

**Writing – original draft:** MEAM, JRB, SAZ, SBB.

**Writing – review & editing:** All authors.

## Competing interests

The authors have declared that no competing interests exist.

## Funding

The authors received no specific funding for this work.

## Data availability

All relevant data are contained within the manuscript and its Supporting Information files. The de-identified individual-level dataset is available from the corresponding author upon reasonable request, subject to institutional data-sharing policies designed to protect patient confidentiality.

## Supporting information

**S1 Checklist.** STROBE checklist for cross-sectional studies.

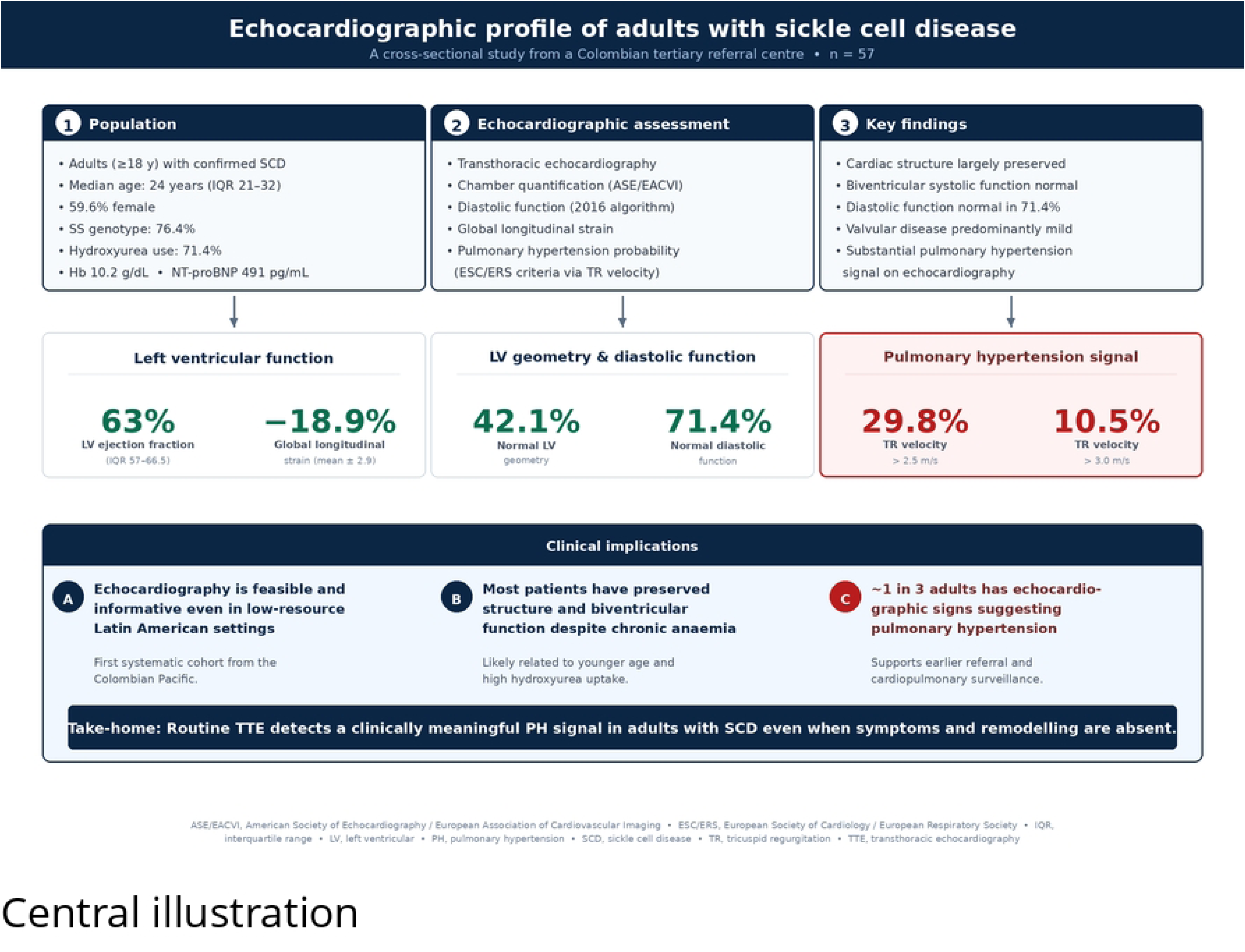

## Notes

### Competing Interest Statement

The authors have declared no competing interest.

### Funding Statement

The author(s) received no specific funding for this work.

### Author Declarations

This retrospective cross-sectional study was approved by the Research Ethics Committee of the Universidad del Valle, Cali, Colombia. The study was classified as risk-free research according to Resolution 8430 of 1993 of the Colombian Ministry of Health, which regulates research with human subjects in Colombia. The requirement for individual written informed consent was waived by the same Ethics Committee given the retrospective design and the use of fully anonymised data extracted from electronic medical records and routine clinical echocardiographic reports. All procedures were conducted in accordance with the Declaration of Helsinki. Patient identifiers were removed before data extraction and analysis.

## References

1. Piel FB, Steinberg MH, Rees DC. Sickle cell disease. N Engl J Med. 2017;376(16):1561–1573.

2. Kato GJ, Piel FB, Reid CD, Gaston MH, Ohene-Frempong K, Krishnamurti L, et al. Sickle cell disease. Nat Rev Dis Primers. 2018;4:18010.

3. Thomson AM, McHugh TA, Oron AP, Teply C, Lonberg N, Tella VV, et al. Global, regional, and national prevalence and mortality burden of sickle cell disease, 2000–2021: a systematic analysis from the Global Burden of Disease Study 2021. Lancet Haematol. 2023;10(8):e585–e599.

4. Ramírez-Cheyne J, Moreno M, Mosquera S, Duque S, Holguín J, Camacho A, et al. First two years of notification of orphan-rare diseases in Cali and identification of some variables associated with mortality. Iatreia. 2020;33(2):111–122.

5. Fitzhugh CD, Lauder N, Jonassaint JC, Telen MJ, Zhao X, Wright EC, et al. Cardiopulmonary complications leading to premature deaths in adult patients with sickle cell disease. Am J Hematol. 2010;85(1):36–40.

6. Hammoudi N, Lionnet F, Redheuil A, Montalescot G. Cardiovascular manifestations of sickle cell disease. Eur Heart J. 2020;41(13):1365–1373.

7. Gladwin MT, Sachdev V. Cardiovascular abnormalities in sickle cell disease. J Am Coll Cardiol. 2012;59(13):1123–1133.

8. Parent F, Bachir D, Inamo J, Lionnet F, Driss F, Loko G, et al. A hemodynamic study of pulmonary hypertension in sickle cell disease. N Engl J Med. 2011;365(1):44–53.

9. Gladwin MT. Cardiovascular complications in patients with sickle cell disease. Hematology Am Soc Hematol Educ Program. 2017;2017(1):423–430.

10. Sachdev V, Machado RF, Shizukuda Y, Rao YN, Sidenko S, Ernst I, et al. Diastolic dysfunction is an independent risk factor for death in patients with sickle cell disease. J Am Coll Cardiol. 2007;49(4):472–479.

11. Abdul-Mohsen MF. Echocardiographic evaluation of left ventricular diastolic and systolic function in Saudi patients with sickle cell disease. J Saudi Heart Assoc. 2012;24(4):217–224.

12. Bedirian R, Soares AR, Maioli MC, de Medeiros JFF, Lopes AJ, Castier MB. Left ventricular structural and functional changes evaluated by echocardiography and two-dimensional strain in patients with sickle cell disease. Arch Med Sci. 2018;14(3):493–499.

13. Naoman SG, Nouraie M, Castro OL, Nwokolo C, Fadojutimi-Akinsiku M, Diaz S, et al. Echocardiographic findings in patients with sickle cell disease. Ann Hematol. 2010;89(1):61–66.

14. Chiadika S, Lim-Fung M, Llanos-Chea F, Canache AS, Yang W, Paruthi C, et al. Echocardiographic parameters to identify sickle cell patients with cardiopathology. Echocardiography. 2018;35(9):1271–1276.

15. von Elm E, Altman DG, Egger M, Pocock SJ, Gøtzsche PC, Vandenbroucke JP, et al. The Strengthening the Reporting of Observational Studies in Epidemiology (STROBE) Statement: guidelines for reporting observational studies. Int J Surg. 2014;12(12):1495–1499.

16. Lang RM, Badano LP, Mor-Avi V, Afilalo J, Armstrong A, Ernande L, et al. Recommendations for cardiac chamber quantification by echocardiography in adults: an update from the American Society of Echocardiography and the European Association of Cardiovascular Imaging. J Am Soc Echocardiogr. 2015;28(1):1–39.e14.

17. Nagueh SF, Smiseth OA, Appleton CP, Byrd BF 3rd, Dokainish H, Edvardsen T, et al. Recommendations for the evaluation of left ventricular diastolic function by echocardiography: an update from the American Society of Echocardiography and the European Association of Cardiovascular Imaging. J Am Soc Echocardiogr. 2016;29(4):277–314.

18. Humbert M, Kovacs G, Hoeper MM, Badagliacca R, Berger RMF, Brida M, et al. 2022 ESC/ERS Guidelines for the diagnosis and treatment of pulmonary hypertension. Eur Heart J. 2022;43(38):3618–3731.

19. Sachdev V, Kato GJ, Gibbs JSR, Barst RJ, Machado RF, Nouraie M, et al. Echocardiographic markers of elevated pulmonary pressure and left ventricular diastolic dysfunction are associated with exercise intolerance in adults and adolescents with homozygous sickle cell anemia in the United States and United Kingdom. Circulation. 2011;124(13):1452–1460.

20. Niihara Y, Miller ST, Kanter J, Lanzkron S, Smith WR, Hsu LL, et al. A phase 3 trial of l-glutamine in sickle cell disease. N Engl J Med. 2018;379(3):226–235.

21. Gordeuk VR, Castro OL, Machado RF. Pathophysiology and treatment of pulmonary hypertension in sickle cell disease. Blood. 2016;127(7):820–828.

22. Mehari A, Gladwin MT, Tian X, Machado RF, Kato GJ. Mortality in adults with sickle cell disease and pulmonary hypertension. JAMA. 2012;307(12):1254–1256.

